# Predicting Mechanical Ventilation Requirement in Guillain–Barré Syndrome using a Multi-Functional Machine Learning Algorithm

**DOI:** 10.64898/2026.06.29.26356838

**Authors:** Yousef Younis, Junshuang Guo

**Affiliations:** Department of Biomedical Engineering, College of Engineering, University of South Florida, Tampa, Florida, USA; Neuro-Intensive Care Unit, First Affiliated Hospital of Zhengzhou University, Zhengzhou, Henan Province, China

**Keywords:** Guillain–Barré syndrome, Mechanical ventilation, Prognosis, Biomarkers, Machine learning

## Abstract

**Background and Aims:** To develop and validate multiple Machine Learning (ML) algorithms that predict Mechanical Ventilation (MV) requirement in Guillain–Barré syndrome (GBS), and to determine whether they outperform the additive, score-based prognostic models in current use.

**Methods:** This retrospective study analysed 233 GBS patients (training set, *n* = 186; validation set, *n* = 47). Five algorithms (Deep Neural Network (DNN), Extreme Gradient Boosting (XGBoost), Logistic Regression (LR), Random Forest (RF), and Naïve Bayes (NB)) were trained and compared. Predictors were chosen by a three-method consensus pipeline executed inside each nested cross-validation fold, retaining 11 features. Whether BorderlineSMOTE was applied was determined per model by Optuna hyperparameter tuning. Hyper-parameter tuning, probability calibration, and bootstrap resampling were applied; performance was evaluated using recall, F1, specificity, AUROC, and Brier score, with SHAP for model interpretability.

**Results:** XGBoost achieved the strongest clinical performance (AUROC 0.807 and recall 0.857), exceeding the validated EGRIS for MV (AUROC *≈*0.62). Sigmoid calibration preserved recall, shifted the operating point by one false positive, and improved the Brier score from 0.210 to 0.110 (BSS *≈* 0.134), so the deployed tool was developed using the probabilities from the calibrated XGBoost model. Consensus selection retained eleven predictors; blood prealbumin, blood FT3, and NLR ranked highest by both embedded importance and SHAP. The model was deployed as an interactive prognostic tool predicting MV risk at admission.

**Interpretation:** ML algorithms substantially improve GBS prognosis by integrating eleven biomarker predictors, modelling nonlinear relationships, and providing SHAP-based interpretability. The single-centre sample is small, so external validation in larger, multi-centre cohorts is required before clinical deployment.

## Introduction

### Context

Guillain–Barré syndrome (GBS) is a severe, acute autoimmune peripheral polyradiculoneuropathy in which demyelination of peripheral nerves causes paralysis that begins distally and progresses proximally.^1–3^ The most common cause of GBS is *Campylobacter jejuni* (*C. jejuni*) infection, which triggers molecular mimicry between bacterial antigens and components of peripheral nerves.^4–6^ Roughly 20–30% of patients require mechanical ventilation (MV) due to respiratory muscle failure,^1,2^ and disease nadir typically occurs 2–4 weeks after onset.^2,3^ Early risk stratification is complicated due to the difficulty of distinguishing GBS variants electrodiagnostically (the current gold standard) during the first weeks after onset. During this period, nerve conduction findings fail to differentiate variants despite their differing outcomes.^2,25^ Because disease nadir falls within this window (2–4 weeks), clinicians must commit to treatment intensity before the outcome of the patient can be confirmed. The current standards of treatment are Intravenous Immunoglobulin (IVIG) and Plasma Exchange (PE); however, identifying patients highest at risk of respiratory failure is crucial to optimize patient-specific treatment and minimize irreversible peripheral nerve damage.^7,8^

### Current prognostic models

There are currently two standard prognostic models: the modified Erasmus GBS Outcome Score (mEGOS), which predicts the inability to walk independently at 4 and 26 weeks, and the Erasmus GBS Respiratory Insufficiency Score (EGRIS), which predicts respiratory insufficiency within the first week of admission. Both models were developed in large prospective cohorts such as the International GBS Outcome Study (IGOS, ∼1,500 patients) and use a simple additive scoring system^9,10^ built on three clinical predictors. Because the day-7 version of mEGOS requires a one-week waiting period, its use is limited at the time when treatment intensity should be decided. Performance of both models is moderate; mEGOS reached an AUROC of ≈ 0.69 for its disability outcome when validated outside its development population,^11^ with comparable performance in pediatric cohorts,^12,13^ and EGRIS yielded an AUROC ≈ 0.62 for MV prediction on external validation in a Chinese cohort.^27^ Both also require regional recalibration when applied outside their development population,^9,11^ limiting portability and deployment. Their simple additive architecture combines only three linearly weighted variables, structurally limiting their ability to capture the complex, nonlinear interactions that occur in the body, interactions that multi-biomarker data can represent.

### Research in biomarkers and machine learning

Recent research demonstrates that biomarkers may provide a more accurate representation of a patient’s physiological state and more prognostic information compared to clinical variables alone.^16–18^ In GBS, the neutrophil-to-lymphocyte ratio (NLR) has been linked to short-term outcome,^24^ cerebrospinal fluid (CSF) and serum neurofilament light chain (NfL) to walking ability,^19,20^ and a range of clinical and electrophysiological variables to respiratory failure.^21^ Machine Learning (ML) models have been shown to be well suited to exploit this information because they can model nonlinear interactions across a large feature dataset, which additive scoring systems cannot.^14,15,26^ Within GBS, Guo et al.^28^ built an interpretable ML pipeline for short-term prognosis (XGBoost AUROC = 0.874 on internal testing); however, that study treats MV as a predictor of prognosis rather than the outcome itself, targeting a binary “good” versus “bad” prognosis defined by the Hughes Functional Grading Scale (HFGS), with good prognosis corresponding to a score of < 4 and bad prognosis to a score of ≥ 4. Comparable pipelines in adjacent autoimmune neurological diseases reach the same conclusion, finding that a nonlinear architecture integrating lab and clinical features outperforms linear scoring systems.^29^

### Goal of this study

This study develops and validates five ML algorithms (DNN, XGBoost, LR, RF, and NB) for MV prediction in GBS, targeting the limitations of existing models. This study uses a multidimensional biomarker feature set, models non-linear interactions, applies SHapley Additive exPlanations (SHAP) at both cohort and individual levels for interpretability, prioritizes minimizing false negatives, and deploys the best-performing model to develop a real-time interactive clinical tool which clinicians can use that predicts outcome at admission.

## Materials and Methods

### Data collection and preprocessing

This retrospective study began with 278 GBS patients admitted to the First Affiliated Hospital of Zhengzhou University. After exclusion of 45 ineligible cases (24 aged <16 years; 21 with a hospital stay <7 days), 233 patients remained. The outcome was MV requirement during hospitalisation. Of the 233 patients, 199 (85.4%) did not require MV and 34 (14.6%) did, yielding an imbalance ratio of 5.85:1. A stratified 80/20 split produced a training set (*n* = 186; 159 non-MV, 27 MV-positive) and a validation set (*n* = 47; 40 non-MV, 7 MV-positive), preserving the class distribution.

Data preprocessing followed a multistage comprehensive pipeline. All symbolic characters were converted to quantitative values. Missing values were imputed using Multiple Imputation by Chained Equations (MICE), which preserves the joint feature distribution (652 missing values across 38 columns). Next, features were normalised to standardised units and three biological ratio predictors were then derived from existing variables in the dataset:^24^

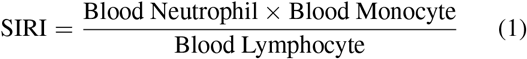

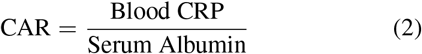

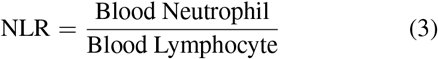

To eliminate data redundancy, individual cell counts were dropped once the ratios were derived, so that the same biological signal was not encoded twice. Finally, features with a Variance Inflation Factor (VIF) >20 were removed to reduce multicollinearity; this threshold (rather than the stricter 10) kept informative features.

### Feature selection

To reduce risk of overfitting the models, predictors were chosen by a three-method consensus framework. Each feature was scored three ways: (i) a *filter test* (Mann–Whitney U for continuous variables, chi-squared for categorical variables, *α* = 0.05); (ii) *bootstrap-stabilised model embedded importance* (30 resamples, selection frequency ≥50%); and (iii) *mutual information* (MI; *k*-NN estimator, *k* = 5, on z-score normalised features, thresholded at the median). Model-embedded importance is derived from each algorithm’s own trained model (e.g., RF embedded importance for the Random Forest algorithm and XGBoost embedded importance for the XGBoost algorithm). Features were z-score normalized for the MI step only (*k*-NN distances are scale-sensitive); XGBoost, being scale-invariant, was trained on raw values, preserving the interpretability of its SHAP split thresholds.

A feature was determined significant enough if it had ≥2/3 votes; complete consensus was not required, because the three methods capture different kinds of relevance, and demanding 3/3 would have removed biologically important features such as albumin quotient and CSF monocyte percentage. Model mean importance broke ties, and the set was capped at 15 features.

To prevent data leakage, the entire pipeline was executed independently inside each fold of a nested 5-fold stratified cross-validation (CV), and a feature was retained only if selected in ≥4/5 folds; the 4/5 threshold (instead of 5/5 folds) tolerates the fold-to-fold fluctuations expected with only 27 MV-positive training patients.

### Model development

To compare different modeling approaches, five different algorithms were trained: Logistic Regression (LR), Naïve Bayes (NB), Random Forest (RF), Extreme Gradient Boosting (XGBoost), and a Deep Neural Network (DNN). Whether BorderlineSMOTE was applied was treated as a hyperparameter and determined per model by Optuna. NB’s minimum recall constraint during threshold selection was relaxed to 0.50 (versus 0.70 for the other models): at 0.70 its threshold collapses toward zero. This is because of its overconfident probability distribution, making both accuracy and AUROC fall. A recall constraint of 0.50 recovers its natural high-confidence operating point (threshold = 0.9999, AUROC = 0.761).

Optuna optimisation (*n*_trials_ = 150) maximised a composite objective:

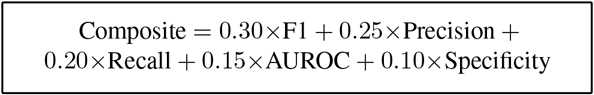

Recall is weighted below precision because it is protected by a minimum constraint (≥0.70) during threshold selection. This approach optimizes performance within that recall floor rather than maximizing it unconditionally, preventing the threshold from collapsing toward zero and blindly flagging every patient as MV-positive. Probability calibration was then applied via sigmoid scaling with 5-fold CV.

Classification thresholds were selected using a leakage-free out-of-fold (OOF) procedure. For each model, stratified 5-fold CV was run on the training data; within each fold, SMOTE (if selected) was applied to the training portion only, so synthetic samples never appeared in a validation fold. The predicted probabilities for each training sample, obtained only when that sample was held out, were pooled across folds. The threshold that maximised the composite operating-point score subject to a minimum-recall constraint of ≥0.70 was selected from these pooled OOF probabilities, then frozen and applied directly to the held-out validation set. Reported operating-point metrics therefore reflect a threshold chosen without any information from the validation data.

### Model evaluation

Model metrics were computed on the held-out validation set, and the Brier score quantified calibration quality against the naïve reference of outcome prevalence. Because AUROC measures rank-order discrimination while Brier score measures probability magnitude accuracy, a model can rank patients well yet remain systematically overconfident (Brier > naïve baseline). Bias-corrected and accelerated (BCa) bootstrap resampling (*n* = 2000) was implemented for confidence intervals, correcting for metric bounding and positive class skewness.

### Model interpretability

SHAP values were computed on the optimized, uncalibrated model^28,29^ using TreeExplainer. Probability calibration is applied as a separate five-fold sigmoid wrapper around clones of the same base classifier and rescales only the model’s output probabilities; it does not alter the feature attributions of the underlying tree ensemble. SHAP attributions were therefore taken from the base classifier, whose feature ranking reflects the model deployed in the interactive tool. SHAP was examined individually via force plots for representative patients.

### Interactive clinical tool

The best-performing model was deployed as an interactive web-based prognostic tool, allowing clinicians to enter patient-specific clinical data and receive a real-time, MV risk probability.

### Statistical analysis

Analyses were performed in Python using scikit-learn, SHAP, Optuna, and dedicated model-specific libraries, such as XGBoost. Continuous variables were compared using the Mann–Whitney U test; categorical variables using the chi-squared or Fisher’s exact test. Significance was set at *P* < 0.05.

## Results

### Study population and data preprocessing

The original dataset contained 278 patients. After the exclusions described in the Methods, 233 patients remained (199 non-MV, 34 MV). After MICE imputation, the final dataset contained no missing values (652 missing entries across 38 columns in the original). Exclusion of the younger patients shifted the age distribution upward as expected, with the mean age rising from 47.5 to 51.4 years and the median from 51.0 to 54.0 years.

Baseline demographic, clinical, and laboratory characteristics of the full cohort, stratified by MV outcome, are presented in Table 1. MV-requiring patients had markedly higher inflammatory indices (NLR, SIRI, CRP, CAR; all *p* ≤ 0.005), lower blood prealbumin (*p* < 0.001) and FT3 (*p* = 0.012), lower CSF monocyte percentage (*p* = 0.021) and albumin quotient (*p* = 0.047), and longer hospital stays (*p* = 0.022), consistent with greater disease severity. Progressive onset was also more common among MV patients compared to non-MV patients (79.4% versus 40.7%, *p* < 0.001), as was PE use (29.4% versus 9.5%, *p* = 0.003).

**Table 1.** Demographic, clinical, and laboratory characteristics of the study population stratified by mechanical ventilation requirement. Univariate analysis of patients.

| Characteristic | Total (n = 233) | No MV (n = 199) | MV Required (n = 34) | p-value |
| --- | --- | --- | --- | --- |
| *Demographics* |  |  |  |  |
| Age (years) | 54.0 (38.0–66.0) | 54.0 (38.0–64.5) | 53.0 (37.5–67.0) | 0.809 |
| Hospitalization days | 14.0 (10.0–22.0) | 13.0 (10.0–21.0) | 18.5 (12.0–27.8) | 0.022 |
| Male sex, n (%) | 135 (57.9) | 118 (59.3) | 17 (50.0) | 0.408 |
| *Clinical features* |  |  |  |  |
| Hughes score at discharge | 3.0 (3.0–4.0) | 3.0 (3.0–4.0) | 4.0 (4.0–5.0) | < 0.001 |
| Progressive onset (PO), n (%) | 108 (46.4) | 81 (40.7) | 27 (79.4) | < 0.001 |
| Hyporeflexia, n (%) | 186 (79.8) | 156 (78.4) | 30 (88.2) | 0.249 |
| Cranial nerve involvement, n (%) | 43 (18.5) | 34 (17.1) | 9 (26.5) | 0.287 |
| Diarrhea, n (%) | 46 (19.7) | 38 (19.1) | 8 (23.5) | 0.714 |
| Upper respiratory infection, n (%) | 29 (12.4) | 24 (12.1) | 5 (14.7) | 0.880 |
| *Laboratory findings* |  |  |  |  |
| Blood WBC ($10^9/L$) | 7.4 (5.7–9.0) | 7.2 (5.6–8.7) | 9.8 (7.2–13.7) | < 0.001 |
| Blood glucose (mg/dL) | 100.2 (85.1–124.5) | 97.3 (83.0–122.1) | 111.1 (99.9–139.2) | 0.004 |
| Blood albumin (mg/dL) | 4110 (3830–4420) | 4150 (3855–4438) | 3965 (3553–4235) | 0.020 |
| Blood prealbumin (mg/dL) | 24.3 (20.7–28.1) | 25.1 (21.3–28.6) | 20.3 (16.8–22.9) | < 0.001 |
| Blood CRP (mg/dL) | 0.2 (0.1–0.8) | 0.2 (0.1–0.7) | 0.7 (0.3–3.2) | < 0.001 |
| Blood FT3 (pg/mL) | 3.0 (2.7–3.3) | 3.0 (2.7–3.3) | 2.8 (2.4–3.0) | 0.012 |
| Blood FT4 (ng/dL) | 1.0 (0.9–1.1) | 1.0 (0.9–1.1) | 1.0 (0.9–1.2) | 0.753 |
| Blood TSH (uIU/mL) | 2.1 (1.3–3.0) | 2.1 (1.4–3.2) | 1.3 (0.6–2.3) | 0.002 |
| *CSF findings* |  |  |  |  |
| CSF leucocyte ($10^6/L$) | 2.0 (2.0–4.0) | 2.0 (2.0–4.3) | 2.0 (2.0–4.0) | 0.369 |
| CSF lymphocyte (%) | 68.0 (60.0–75.0) | 67.0 (60.0–73.8) | 72.8 (61.0–79.7) | 0.020 |
| CSF monocyte (%) | 30.0 (23.0–37.0) | 31.0 (24.0–38.0) | 23.7 (16.2–35.5) | 0.021 |
| CSF protein (mg/dL) | 66.3 (41.6–101.5) | 67.2 (43.3–101.8) | 46.5 (34.5–93.2) | 0.055 |
| CSF glucose (mmol/L) | 3.9 (3.3–4.9) | 3.8 (3.3–4.8) | 4.3 (3.6–5.5) | 0.060 |
| Albumin quotient ($10^{-3}$) | 10.2 (5.7–17.1) | 10.7 (6.0–17.0) | 6.4 (4.3–17.1) | 0.047 |
| IgG index | 0.8 (0.6–0.9) | 0.8 (0.6–0.9) | 0.9 (0.5–1.0) | 0.236 |
| *Derived inflammatory indices* |  |  |  |  |
| NLR | 3.1 (1.9–5.5) | 2.8 (1.9–4.9) | 7.0 (4.2–15.2) | < 0.001 |
| SIRI | 1.5 (0.8–3.1) | 1.3 (0.8–2.6) | 4.2 (1.9–7.7) | < 0.001 |
| CAR ($10^{-4}$) | 0.4 (0.1–1.5) | 0.3 (0.1–1.2) | 1.0 (0.3–2.0) | 0.005 |
| *Treatment* |  |  |  |  |
| IVIG, n (%) | 191 (82.0) | 162 (81.4) | 29 (85.3) | 0.761 |
| PE, n (%) | 29 (12.4) | 19 (9.5) | 10 (29.4) | 0.003 |
| GCS treatment, n (%) | 44 (18.9) | 34 (17.1) | 10 (29.4) | 0.144 |

MV, mechanical ventilation; WBC, white blood cell count; CRP, C-reactive protein; FT3, free triiodothyronine; FT4, free thyroxine; TSH, thyroid-stimulating hormone; CSF, cerebrospinal fluid; NLR, neutrophil-to-lymphocyte ratio; SIRI, systemic inflammatory response index; CAR, C-reactive protein-to-albumin ratio; IVIG, intravenous immunoglobulin; PE, plasma exchange; GCS, glucocorticosteroid.

### Feature selection results

The three-method consensus pipeline identified eleven features that met the stability criterion of selection in at least four of five folds: blood prealbumin, blood CRP, blood albumin, CAR, blood FT3, SIRI, NLR, albumin quotient, CSF monocyte percentage, IgG index, and blood glucose. Six of the eleven were endorsed by all three methods; the remaining five received two votes but still satisfied the fold-stability criterion (≥4/5 folds). The consensus voting structure is summarised in Table 2.

**Table 2.** Feature importance ranking and consensus selection results for mechanical ventilation prediction in GBS (training set: No MV *n* = 159, MV-required *n* = 27). Features are ordered by bootstrapped XGBoost embedded importance score (*n* = 30 iterations), the primary ranking criterion.

| Feature | No MV (mean±SD) | MV Req. (mean±SD) | $p$-value | Stat. | XGB | MI | Votes | Importance | CV Stab. |
| --- | --- | --- | --- | --- | --- | --- | --- | --- | --- |
| Blood Prealbumin (mg/dL) | 25.23 ± 6.17 | 18.99 ± 4.89 | < 0.001 | 1 | 1 | 1 | 3/3 | 0.103 | 5/5 |
| Blood FT3 (pg/mL)<sup>†</sup> | 3.74 ± 4.43 | 2.83 ± 1.11 | 0.002 | 1 | 1 | 1 | 3/3 | 0.052 | 5/5 |
| NLR | 4.28 ± 4.25 | 15.51 ± 34.48 | < 0.001 | 1 | 1 | 1 | 3/3 | 0.051 | 5/5 |
| SIRI | 2.46 ± 2.98 | 12.89 ± 38.73 | < 0.001 | 1 | 1 | 1 | 3/3 | 0.042 | 5/5 |
| Blood CRP (mg/dL) | 1.29 ± 4.23 | 2.28 ± 2.98 | < 0.001 | 1 | 1 | 1 | 3/3 | 0.042 | 5/5 |
| Blood glucose (mg/dL) | 108.87 ± 41.12 | 126.06 ± 45.99 | 0.013 | 1 | 1 | 0 | 2/3 | 0.038 | 4/5 |
| CAR(10<sup>-4</sup>) | 2.91 ± 11.21 | 3.51 ± 7.10 | 0.014 | 1 | 1 | 0 | 2/3 | 0.036 | 4/5 |
| Albumin quotient (10<sup>-3</sup>) | 14.23 ± 11.95 | 14.22 ± 18.36 | 0.071 | 0 | 1 | 1 | 2/3 | 0.031 | 4/5 |
| CSF Monocyte (%) | 30.75 ± 10.09 | 28.63 ± 15.72 | 0.119 | 0 | 1 | 1 | 2/3 | 0.030 | 4/5 |
| Blood Albumin (mg/dL) | 4093.06 ± 514.82 | 3804.26 ± 544.87 | 0.003 | 1 | 1 | 1 | 3/3 | 0.029 | 5/5 |
| IgG index | 0.86 ± 1.13 | 0.84 ± 0.37 | 0.486 | 0 | 1 | 1 | 2/3 | 0.023 | 5/5 |

1 = endorsed by this method; 0 = not endorsed. NLR, neutrophil-to-lymphocyte ratio; SIRI, systemic inflammatory response index; CRP, C-reactive protein; FT3, free triiodothyronine; CAR, C-reactive protein-to-albumin ratio; CSF, cerebrospinal fluid. <sup>†</sup>The large No-MV SD for FT3 (4.43 pg/mL, exceeding the mean of 3.74 pg/mL) is driven by outliers; the IQR (2.70–3.38 pg/mL) is consistent with the normal physiological range and is the robust summary.

Notably, the albumin quotient (*p* = 0.071), CSF monocyte percentage (*p* = 0.119), and IgG index (*p* = 0.486) all failed the Mann–Whitney test, yet were retained by the consensus pipeline.

Features are ranked in Table 2 by XGBoost embedded importance, the primary ranking criterion. Blood prealbumin was the most influential predictor (importance 0.103), followed by the remaining predictors listed in Table 2.

### Machine learning model performance

#### i. Internal cross-validation

Nested five-fold CV for XGBoost, with feature selection performed inside each training fold to prevent leakage, yielded mean ROC-AUC 0.765 ± 0.075, PR-AUC 0.502 ± 0.150, recall 0.813 ± 0.018, and F1 0.470 ± 0.081. The narrow recall SD reflects discrete increments of approximately 0.18 per misclassified patient with ∼5–6 MV-positive cases per validation fold. The five fold-level recalls were [0.833, 0.800, 0.800, 0.800, 0.833].

#### ii. Hold-out test set performance

Table 3 and Figure 1 summarize performance on the validation set (n = 47). During hyperparameter optimization, Optuna found SMOTE did not improve the CV performance for the XGBoost model and thus it was not applied. XGBoost achieved the strongest discrimination (AUROC = 0.807, recall = 0.857), missing only one of 7 MV cases (FN = 1). RF matched FN = 1 but at a lower AUROC (0.732) and at a lower threshold than XGBoost (meaning it required less confidence to flag a patient as needing MV). DNN and NB posted the highest raw accuracies (0.894 and 0.915) but each missed three cases (FN = 3). In this imbalanced setting, high accuracy reflects exploitation of the majority class and is not a reliable indicator of clinical performance. NB maintained good rank-order discrimination (AUROC = 0.761) despite overconfident probabilities (threshold = 0.9999). LR was the weakest discriminator (AUROC = 0.679, FN = 3), failing to capture nonlinear relationships. Because false negatives carry higher clinical cost than false positives during MV risk prediction, XGBoost was selected for deployment.

**Table 3.**
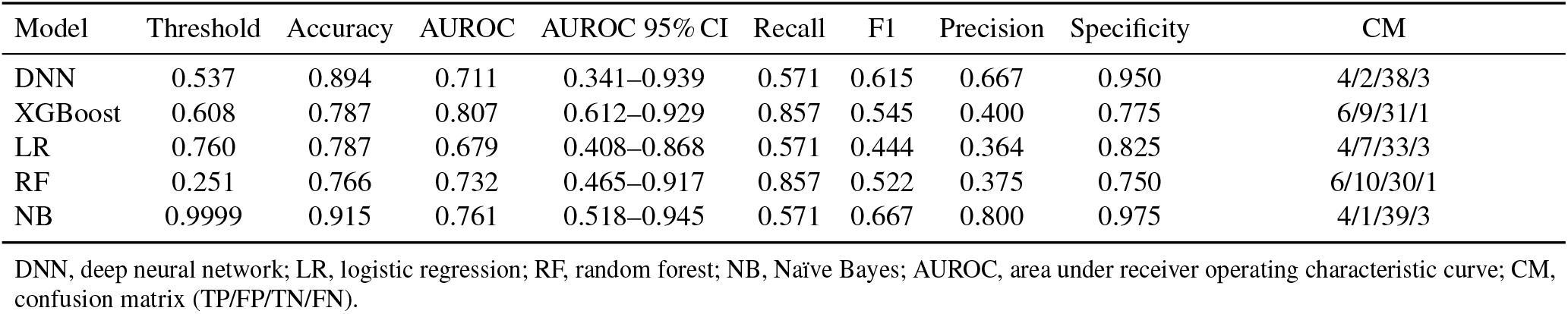
Performance of five machine learning algorithms on the hold-out validation set (*n* = 47; MV+: 7) for mechanical ventilation prediction in Guillain–Barré syndrome (uncalibrated models). Confusion matrix values reported as TP/FP/TN/FN.

| Model | Threshold | Accuracy | AUROC | AUROC 95% CI | Recall | F1 | Precision | Specificity | CM |
| --- | --- | --- | --- | --- | --- | --- | --- | --- | --- |
| DNN | 0.537 | 0.894 | 0.711 | 0.341–0.939 | 0.571 | 0.615 | 0.667 | 0.950 | 4/2/38/3 |
| XGBoost | 0.608 | 0.787 | 0.807 | 0.612–0.929 | 0.857 | 0.545 | 0.400 | 0.775 | 6/9/31/1 |
| LR | 0.760 | 0.787 | 0.679 | 0.408–0.868 | 0.571 | 0.444 | 0.364 | 0.825 | 4/7/33/3 |
| RF | 0.251 | 0.766 | 0.732 | 0.465–0.917 | 0.857 | 0.522 | 0.375 | 0.750 | 6/10/30/1 |
| NB | 0.9999 | 0.915 | 0.761 | 0.518–0.945 | 0.571 | 0.667 | 0.800 | 0.975 | 4/1/39/3 |

DNN, deep neural network; LR, logistic regression; RF, random forest; NB, Naïve Bayes; AUROC, area under receiver operating characteristic curve; CM, confusion matrix (TP/FP/TN/FN).

**Figure 1.**
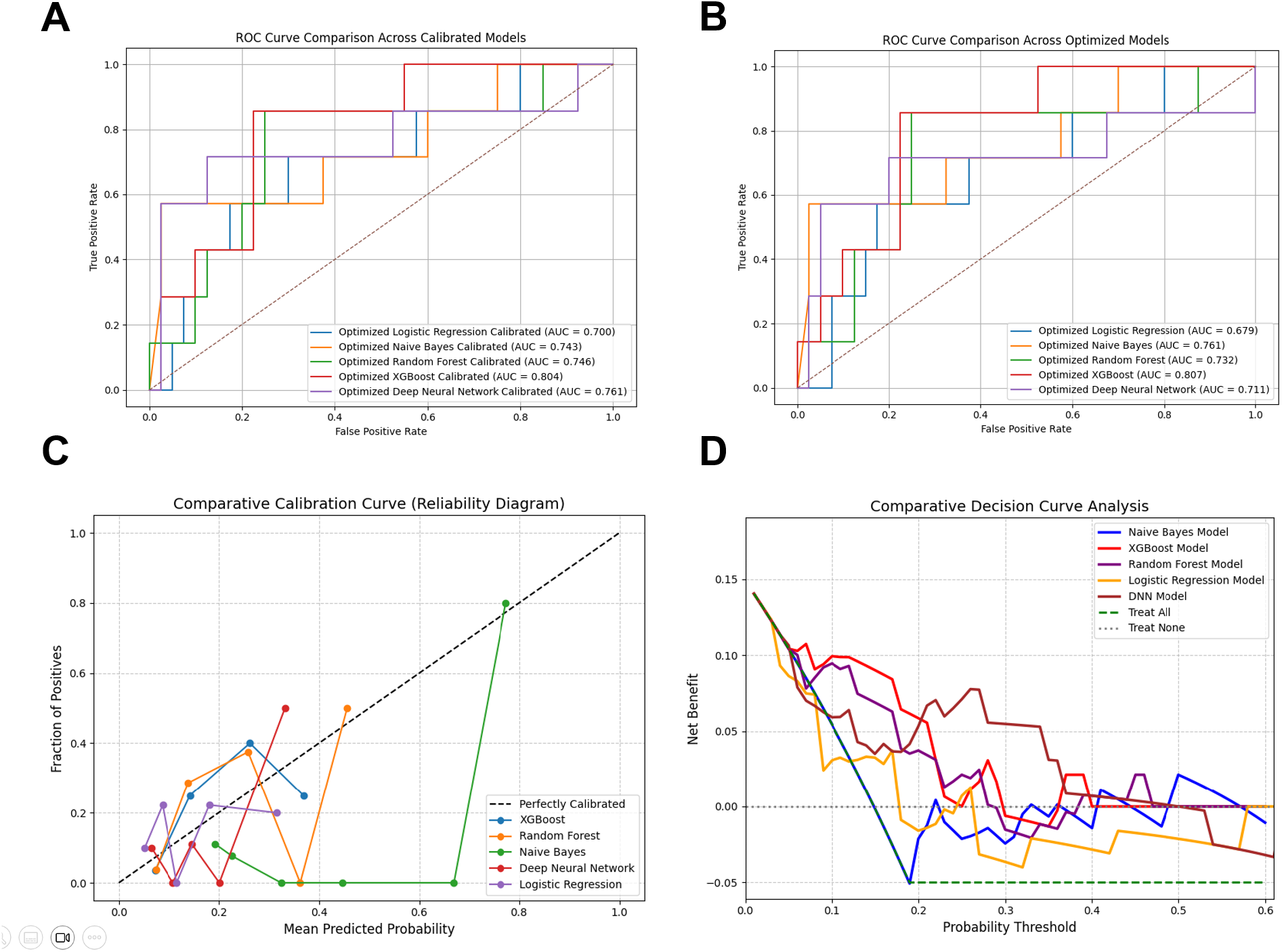
A: ROC curves of all optimised models (uncalibrated); B: ROC curves of all calibrated models; C: Comparative calibration curve (reliability diagram) of all calibrated ML models; D: Comparative decision curve analysis of all calibrated ML models.

#### iii. Probability calibration

Sigmoid calibration (5-fold CV) adjusted predicted probabilities toward empirical outcome frequencies without altering the AUROC (0.807). At the OOF-derived threshold of 0.161, the confusion matrix is 6/10/30/1 (TP/FP/TN/FN), achieving a recall of 0.857 with one additional false positive and no additional false negatives. Calibration thus corrects the probability scale without altering the decision boundary.

The uncalibrated Brier score of 0.210 exceeds the naïve baseline (0.127), confirming overconfidence in the raw outputs. After calibration the Brier score improves to 0.110 (BSS ≈ 0.134). The calibrated model is deployed in the interactive tool. The performance of the uncalibrated and calibrated XGBoost models are reported in Tables 3 and 5, respectively.

**Table 4.** Threshold sensitivity analysis for the calibrated XGBoost model on the hold-out validation set (*n* = 47; MV+: 7). AUROC is threshold-independent and remains constant across all rows at 0.807. The deployed threshold of 0.161 falls between the 0.15^†^ and 0.20^†^ rows.

| Threshold | Sensitivity | Specificity | Precision | F1 | Accuracy | TP/FP/TN/FN |
| --- | --- | --- | --- | --- | --- | --- |
| 0.05 | 1.0000 | 0.0000 | 0.1489 | 0.2593 | 0.1489 | 7/40/0/0 |
| 0.10 | 0.8571 | 0.7000 | 0.3333 | 0.4800 | 0.7234 | 6/12/28/1 |
| **0.15<sup>†</sup>** | **0.8571** | **0.7500** | **0.3750** | **0.5217** | **0.7660** | **6/10/30/1** |
| 0.20 | 0.7143 | 0.7750 | 0.3571 | 0.4762 | 0.7660 | 5/9/31/2 |
| 0.25 | 0.4286 | 0.7750 | 0.2500 | 0.3158 | 0.7234 | 3/9/31/4 |
| 0.30 | 0.1429 | 0.9250 | 0.2500 | 0.1818 | 0.8085 | 1/3/37/6 |
| 0.35 | 0.1429 | 0.9250 | 0.2500 | 0.1818 | 0.8085 | 1/3/37/6 |

Sensitivity = recall = TP / (TP + FN). CM format: TP / FP / TN / FN ( $n_{\text{pos}} = 7$ , $n_{\text{neg}} = 40$ ). AUROC is threshold-independent and constant across rows.
<sup>†</sup>Rows bracketing the deployed threshold of 0.161.

**Table 5.**
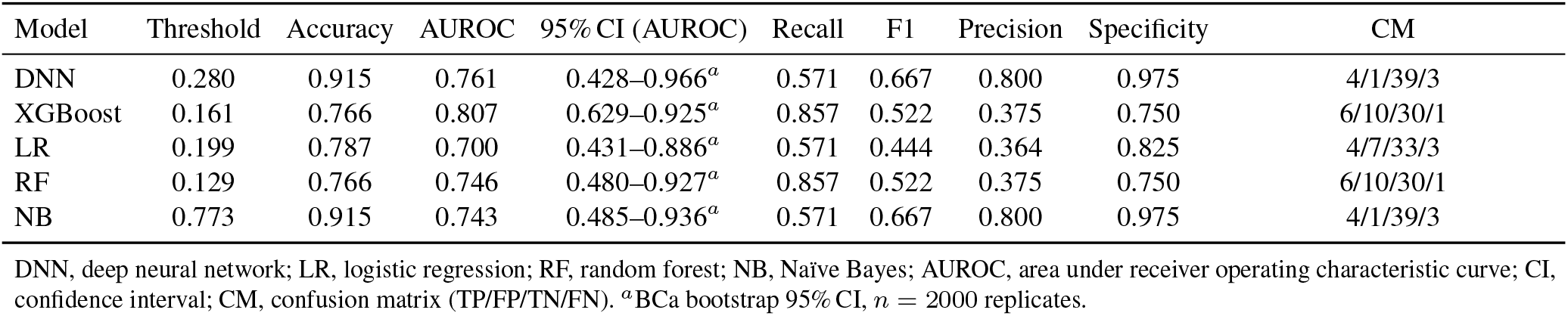
Performance of machine learning algorithms after sigmoid probability calibration (5-fold cross-validation) on the hold-out validation set (*n* = 47; MV+: 7). Confusion matrix values reported as TP/FP/TN/FN.

#### iv. Bootstrap validation

BCa bootstrap resampling (*n* = 2000) of the uncalibrated XGBoost model yielded a resampled-mean accuracy 0.789 (95% CI: 0.681–0.915), AUROC 0.807 (0.612–0.929), recall 0.854 (0.286–1.000), and specificity 0.777 (0.650–0.900). BCa correction was applied because recall has a bounded, skewed distribution with only *n* = 7 positive-class observations, for which standard percentile intervals would produce incorrect results. The wide recall CI reflects the small positive-class sample rather than classifier instability, as a single misclassified patient shifts recall by ∼14.3 points.

#### v. Threshold sensitivity analysis

Table 4 reports the calibrated model’s sensitivity, specificity, precision, F1, and accuracy across thresholds from 0.05 to 0.35 (AUROC is threshold-independent at 0.807). The deployed threshold of 0.161 achieves a recall of 0.857, missing one of seven MV-positive patients. Clinicians wishing to prioritise ruling out MV requirement may lower the threshold at the cost of reduced specificity; for example, lowering the threshold to 0.05 achieves full sensitivity at the expense of zero specificity. Raising the threshold beyond 0.20 reduces recall, and the tradeoff around the deployed point is steep. This range defines the decision boundary clinicians can adjust according to institutional risk tolerance.

### Model interpretability via SHAP

SHAP analysis confirmed blood prealbumin as the most influential predictor, with elevated levels exerting a strong protective effect on predicted MV risk; albumin quotient and SIRI produced the next largest contributions, with CSF monocyte percentage exerting the strongest risk-increasing attribution. The strongest protective interaction was between prealbumin and albumin quotient.

Individual force plots reproduced the multidimensional pattern: elevated prealbumin and albumin quotient drove predictions toward a favourable outcome, whereas reduced CSF monocyte percentage and elevated inflammatory markers pushed predictions toward MV requirement (Figure 2).

**Figure 2.**
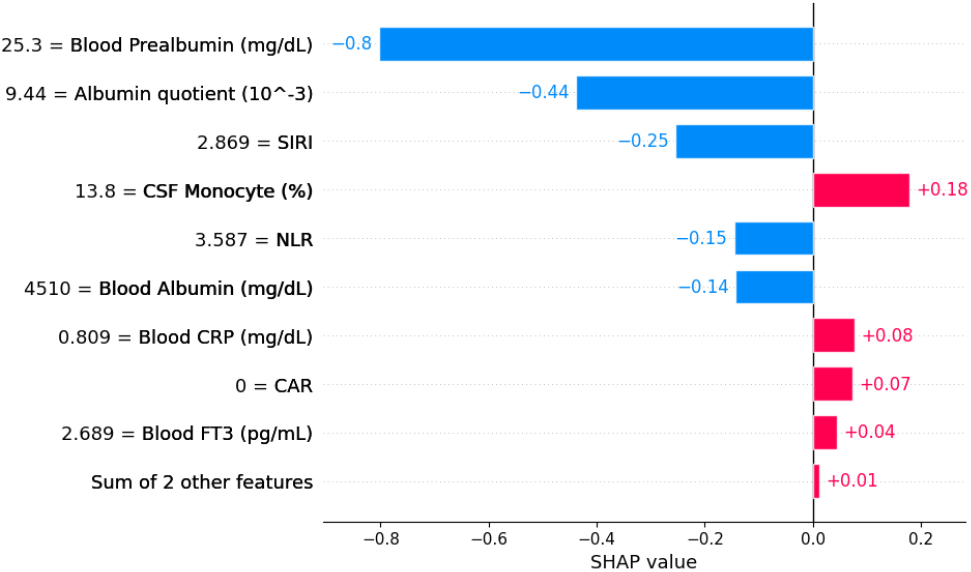
SHAP interpretation of the XGBoost model for a representative high-confidence MV-positive patient. Prealbumin and albumin quotient contribute the largest protective attributions; the directional bar plot shows features driving the prediction toward (positive SHAP) or away from (negative SHAP) MV requirement.

### Interactive prognostic tool

A web-based interactive tool was developed and deployed (Figure 3). Clinicians enter the eleven selected features, and the tool returns the calibrated MV probability with a bootstrap confidence interval, a SHAP-based feature contribution breakdown, and a risk stratification: low (< 0.10), moderate (0.10–0.16), high (> 0.16). A threshold of > 0.16 was reported as high because the calibrated XGBoost model’s threshold is 0.161. Above this threshold, a patient would be flagged as predicted to require MV, and the low-risk boundary of 0.10 marks probabilities below the cohort’s MV prevalence (≈0.15).

**Figure 3.**
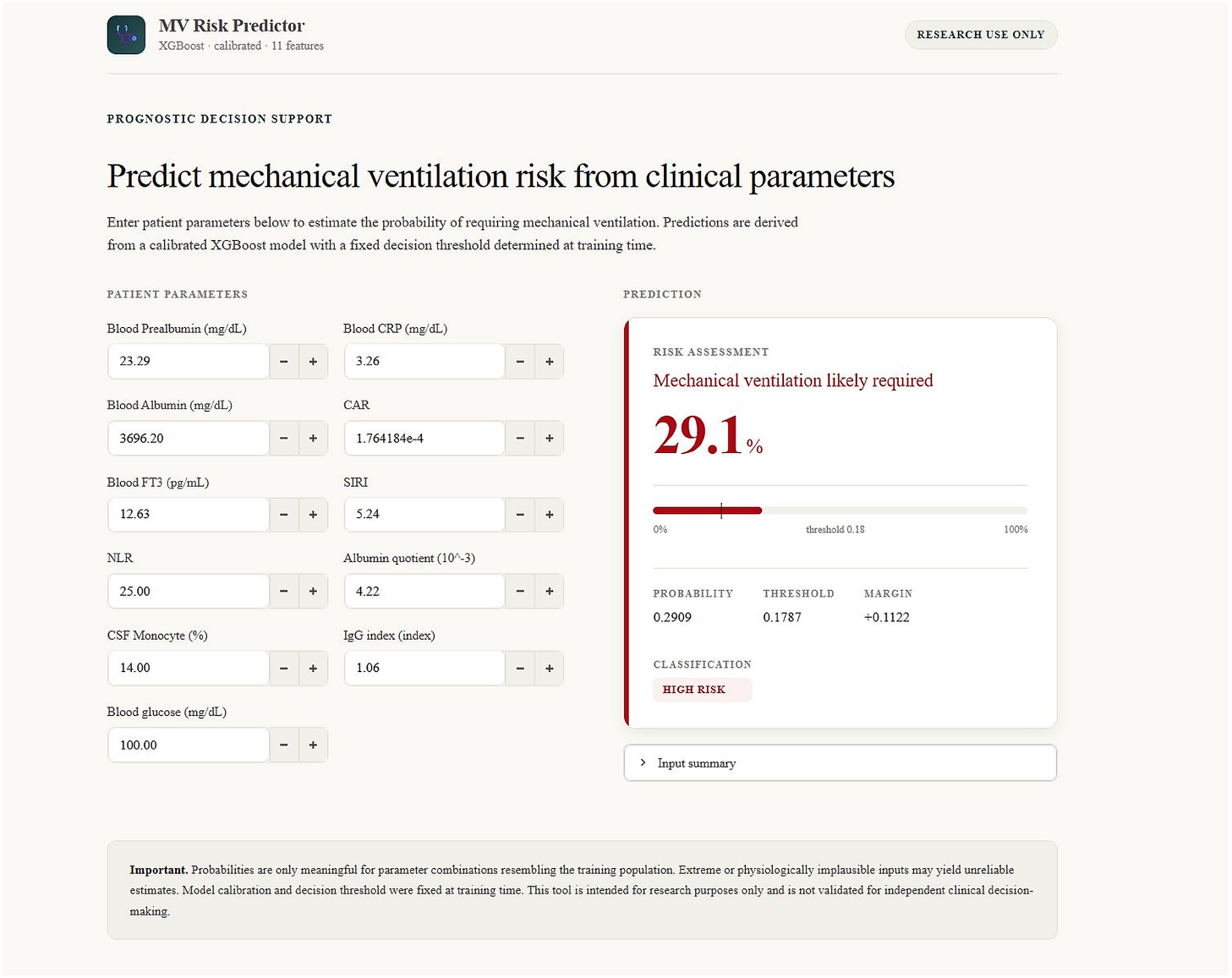
Interactive web-based clinical tool for patient-specific MV risk prediction. The deployed model is the calibrated XGBoost classifier.

## Discussion

XGBoost achieved the strongest clinical performance (AUROC 0.807, 1 FN of 7 MV+ cases), exceeding the externally validated MV performance of EGRIS (≈ 0.62^27^) and the disability-oriented mEGOS (≈ 0.69^11^). This improvement in discriminative power reflects two factors: the use of eleven biomarker predictors rather than three clinical variables, and a machine learning architecture capable of capturing non-linear, interaction-dependent relationships in the data. The leakage-safe, fold-by-fold validation pipeline^15^ ensures that this performance estimate is unbiased rather than inflated by data leakage.

### Clinical significance of key predictors

Blood prealbumin was the strongest predictor across all three selection methods and SHAP. Its short half-life makes it a rapid index of inflammatory status, and its suppression in GBS likely reflects both malnutrition and systemic inflammation.^16,24^ The simultaneous selection of NLR, SIRI, CRP, and CAR indicates that inflammation drives MV risk through multiple complementary, non-redundant axes.^23^ Blood FT3 reflects non-thyroidal illness syndrome, linking thyroid-axis suppression to GBS severity, while CSF monocyte percentage and albumin quotient extend the signal into the CNS compartment, capturing intrathecal immune activity and blood–brain barrier integrity.^16,22^ This diversity of axes (inflammation, nutrition, thyroid function, blood–brain barrier integrity, and intrathecal immunity) explains why a multidimensional ML model outperforms a three-variable additive score, as each carries an independent fragment of the prognostic signal that additive summation cannot integrate.

The SHAP force plot (Figure 2) confirms this multidimensional picture at the individual level. For the highest-confidence true-positive patient, nutritional and CNS-compartment markers dominated the protective attributions, with inflammatory markers contributing in their expected directions. The local attribution magnitudes reflect the specific feature values of that patient rather than the global importance hierarchy, illustrating that the model weights each feature according to the individual’s clinical profile rather than applying a fixed ranking across all patients.

Albumin quotient, CSF monocyte percentage, and IgG index were retained and confirmed informative by SHAP despite failing the univariate filter (*p* = 0.071, 0.119, 0.486), showing that univariate significance does not guarantee multivariate predictive value. Features can appear unimportant in isolation yet contribute meaningfully in combination with others, a pattern marginal-association tests miss but methods such as XGBoost embedded importance and MI can capture.

### Why XGBoost outperformed other algorithms

LR’s weak discrimination is because the most informative predictors (NLR, SIRI, and CAR) are skewed derived ratios with threshold-like rather than linear relationships to outcome, which a linear model cannot capture without explicit interaction terms; its extreme L2 regularisation further compressed raw outputs, explaining the large calibration gain in Brier score (0.497 to 0.124). NB and the DNN share a related failure mode: both yielded high raw accuracy and high FN counts by defaulting to the majority class, NB through overconfident probability estimates and the DNN through insufficient data relative to model capacity, illustrating why AUROC and clinical utility diverge under class imbalance. RF’s lower AUROC than XGBoost despite an identical falsenegative count suggests that gradient boosting with Optuna-tuned regularisation extracted more discriminative signal from the small training set than bagging.

### Calibration and probability communication

The uncalibrated XGBoost Brier score (0.210) exceeds the naïve baseline (0.127), as its raw outputs are well ranked (AUROC = 0.807) but systematically overconfident, because AUROC measures rank order whereas the Brier score measures the accuracy of the probability magnitudes. Sigmoid calibration corrects this (Brier 0.110, BSS ≈ 0.134) without altering recall or the single missed case; the calibrated operating point carries one additional false positive (specificity 0.750 vs. 0.775). The deployed tool therefore reports calibrated probabilities, while cross-algorithm comparison uses the uncalibrated metrics.

### Comparison with prior research

NLR emerged as a top SHAP-ranked predictor both here and in Guo et al.,^28^ despite different outcomes and selection methods, suggesting that systemic inflammatory burden carries a robust signal across multiple GBS outcomes. Blood prealbumin, the strongest predictor identified here, has not featured prominently in prior GBS literature, which has focused on CSF NfL and electrophysiological subtyping;^19,20,25^ its prominence suggests that systemic nutritional and inflammatory status carries MV-specific prognostic information that has been underexploited. Further research is required to explore the role of Blood Prealbumin as a prognostic indicator. The complementary selection of inflammatory markers in Yao et al.,^27^ who also modelled MV in GBS, further supports the central role of inflammatory burden in respiratory deterioration.

The present work also evaluates five algorithm families under identical preprocessing, split, and feature-selection conditions, and reports both AUROC discrimination and Brier calibration, a pairing essential once the output is a patient-specific probability rather than a binary flag, which previous GBS ML studies have not systematically addressed.^15^

### Clinical implications

Like EGRIS, the present tool predicts from admission data, but unlike EGRIS, it integrates eleven biomarkers and provides SHAP-based patient-level explanations that support shared decision-making and reduce the black-box barrier to ML adoption.^14,15^ Additionally, this tool avoids the one-week delay of the day-7 mEGOS. Because BorderlineSMOTE is evaluated during Optuna tuning rather than applied by default, imbalance handling adapts if the model is retrained on a new local dataset, removing the regional recalibration requirement that constrains existing GBS prognostic models. Moreover, if a local dataset of GBS patients is available, the model can be retrained on that cohort to predict the probability of MV risk in patients from the same or similar clinical setting. Despite the methodological rigour of the present study, this tool is intended as a decision support tool, not a standalone decision rule; clinical judgment should determine the final management decision.

## Conclusions

XGBoost substantially improves MV prognosis in GBS over existing scoring systems, integrating eleven biomarker predictors through a leakage-safe consensus pipeline, capturing nonlinear interactions, and explaining each prediction through SHAP. On an independent validation set it achieves AUROC = 0.807 and recall = 0.857 with only one false negative per seven MV-positive patients, above the externally validated EGRIS for MV (AUROC ≈ 0.62) and the disability-oriented mEGOS (≈ 0.69), and sigmoid calibration corrects the raw overconfidence (Brier 0.210 to 0.110, BSS ≈ 0.134) without altering the decision boundary. The deployed tool generates calibrated, patient-specific risk at admission from routine laboratory data, removing the regional recalibration that constrains existing scores. Blood prealbumin, the strongest individual predictor, is underexploited in prior GBS literature and warrants prospective investigation as an actionable nutritional and inflammatory biomarker. External validation in larger, multi-centre cohorts is the essential next step before clinical deployment.

## Limitations

The dataset comprises 233 GBS patients from a single centre; since GBS is rare, larger multi-centre cohorts are required to validate performance across different regions.^21,27^ As a retrospective study, it is also subject to selection and information bias. External validation on independent multicentre data has not yet been performed and is critical before clinical deployment.^15^ The large class imbalance (34 MV-positive vs. 199 non-MV) constrains ML performance, producing wide 95% bootstrap confidence intervals and a low F1 score. Temporal biomarker trajectories across the disease course were not modeled, so the tool predicts whether, not when, a patient will require MV. Finally, the decision curve analysis shows near-zero net benefit at high decision thresholds, consistent with the small positive-class fraction, so the tool, at its current state, should function as a prognostic decision support tool rather than a standalone decision rule, with clinical judgment determining the final management decision.

## Abbreviations

GBS: Guillain–Barré syndrome;
ML: machine learning;
MV: mechanical ventilation
mEGOS: modified Erasmus GBS Outcome Score
EGRIS: Erasmus GBS Respiratory Insufficiency Score
AUROC: area under the receiver operating characteristic curve
AUC: area under the curve
SHAP: SHapley additive exPlanation
DNN: deep neural network
LR: logistic regression
RF: random forest
NB: Naïve Bayes
SMOTE: Synthetic Minority Oversampling Technique
CSF: cerebrospinal fluid
VIF: Variance Inflation Factor
MICE: Multiple Imputation by Chained Equations
SIRI: systemic inflammatory response index
CAR: C-reactive protein to albumin ratio
NLR: neutrophil-to-lymphocyte ratio
PE: plasma exchange
IVIG: intravenous immunoglobulin
NfL: neurofilament light chain
MI: mutual information
CV: cross-validation
CI: confidence interval
BSS: Brier Skill Score.

## Ethics Approval

This study was approved by the Ethics Committee of the First Affiliated Hospital of Zhengzhou University (2022-KY-0176). Informed consent was obtained from all participants.

## Funding

This work received no external funding. Computational resources were provided by Google Colab Pro.

## Data Availability

The datasets used in this study are available from the corresponding author upon reasonable request.

## CRediT Authorship Contribution Statement

Yousef Younis: Conceptualization, Methodology, Software, Formal Analysis, Data Curation, Writing – Original Draft, Visualization. Junshuang Guo: Resources, Investigation, Writing – Review & Editing, Supervision, Validation, Project Administration.

## Declaration of Competing Interest

The authors declare that they have no competing interests.

## Acknowledgements

We acknowledge the clinical staff at the First Affiliated Hospital of Zhengzhou University for patient data collection and clinical support. The structure, framework, and scientific content of this paper were developed by the authors; AI tools were used to improve sentence clarity and conciseness and assist with LaTeX formatting.

## Notes

### Competing Interest Statement

The authors have declared no competing interest.

### Author Declarations

Ethics committee of the First Affiliated Hospital of Zhengzhou University gave ethical approval for this work (approval number 2022-KY-0176). Informed consent was obtained from all participants.

## References

[1] Willison HJ, Jacobs BC, van Doorn PA. Guillain–Barré syndrome. Lancet. 2016;388(10045):717–717. doi:10.1016/S0140-6736(16)00339-1

[2] van den Berg B, et al. Guillain–Barré syndrome: pathogenesis, diagnosis, treatment and prognosis. Nat Rev Neurol. 2014;10(8):469–469. doi:10.1038/nrneurol.2014.121

[3] Bellanti R, Rinaldi S. Guillain–Barré syndrome: a comprehensive review. Eur J Neurol. 2024;31(1):e16365. doi:10.1111/ene.16365

[4] Finsterer J. Triggers of Guillain–Barré syndrome: Campylobacter jejuni predominates. Int J Mol Sci. 2022;23(22):14222. doi:10.3390/ijms232214222

[5] Rojas M, et al. Molecular mimicry and autoimmunity. J Autoimmun. 2018;95:100–123. doi:10.1016/j.jaut.2018.10.012

[6] Laman JD, et al. Guillain–Barré syndrome: expanding the concept of molecular mimicry. Trends Immunol. 2022;43(4):296–296. doi:10.1016/j.it.2022.02.003

[7] Wu X, et al. Predictors for mechanical ventilation and short-term prognosis in patients with Guillain–Barré syndrome. Crit Care. 2015;19(1):310. doi:10.1186/s13054-015-1037-z

[8] Wen P, et al. Risk factors for the severity of Guillain–Barré syndrome. Sci Rep. 2021;11(1). doi:10.1038/s41598-021-91132-3

[9] Doets AY, et al. Predicting outcome in Guillain–Barré syndrome: international validation of the modified Erasmus GBS outcome score. Neurology. 2022;98(5):e518–e532. doi:10.1212/WNL.0000000000013139

[10] Xue G, et al. Construction and evaluation of a prognostic prediction model based on the mEGOS score. Front Neurol. 2023;14:1303243. doi:10.3389/fneur.2023.1303243

[11] Papri N, et al. Validation and adjustment of modified Erasmus GBS outcome score in Bangladesh. Ann Clin Transl Neurol. 2022;9(8):1264–1264. doi:10.1002/acn3.51627

[12] Ulfa M, Widowati T, Triono A. EGOS to predict functional outcomes. Paediatr Indones. 2022;62(2):130–130. doi:10.14238/pi62.2.2022.130-7

[13] Chaweekulrat P, Sanmaneechai O. Prognostic model for time to achieve independent walking in children with GBS. Pediatr Res. 2022;92(5):1417–1417. doi:10.1038/s41390-021-01919-3

[14] Vivas AJ, Boumediene S, Tobón GJ. Predicting autoimmune diseases. Autoimmun Rev. 2024;23(9):103611. doi:10.1016/j.autrev.2024.103611

[15] Diaz-Uriarte R, et al. Ten quick tips for biomarker discovery and validation analyses using machine learning. PLOS Comput Biol. 2022;18(8):e1010357. doi:10.1371/journal.pcbi.1010357

[16] Wang Y, et al. Biomarkers of Guillain–Barré syndrome. Mediators Inflamm. 2015;2015:564098. doi:10.1155/2015/564098

[17] Hansson O. Biomarkers for neurodegenerative diseases. Nat Med. 2021;27(6):954–963. doi:10.1038/s41591-021-01382-x

[18] Khalil M, et al. Neurofilaments as biomarkers in neurological disorders. Nat Rev Neurol. 2024. doi:10.1038/s41582-024-00955-x

[19] Hafsteinsdóttir B, et al. Neurofilament light chain as a diagnostic and prognostic biomarker in GBS. J Neurol. 2024;271(11):7282–7282. doi:10.1007/s00415-024-12679-5

[20] Jin M, et al. CSF neurofilament light chain predicts short-term prognosis in pediatric GBS. Front Neurol. 2022;13:972367. doi:10.3389/fneur.2022.972367

[21] Galassi G, et al. Predictors of respiratory failure in GBS: a 22 year cohort study. Eur J Neurol. 2024;31(1):e16090. doi:10.1111/ene.16090

[22] Li P, et al. Identification of CSF biomarkers by proteomics in GBS. Exp Ther Med. 2018;15(6):5177–5177. doi:10.3892/etm.2018.6117

[23] Wu C-L, et al. Plasma biomarkers reflect immune mechanisms of GBS. Front Neurol. 2021;12:720794. doi:10.3389/fneur.2021.720794

[24] Ethemoglu O, Calik M. Effect of serum inflammatory markers on the prognosis of GBS. Neuropsychiatr Dis Treat. 2018;14:1255–1260. doi:10.2147/NDT.S162896

[25] Arends S, et al. Electrodiagnostic subtyping in GBS in the IGOS. Eur J Neurol. 2024;31(9):e16335. doi:10.1111/ene.16335

[26] Nguyen QTN, et al. Machine learning approaches for predicting 5-year breast cancer survival. Cancer Sci. 2023;114(10):4063–4063. doi:10.1111/cas.15917

[27] Yao J, et al. Predicting of mechanical ventilation and outcomes in GBS. Neurol Ther. 2023;12(6):2121–2121. doi:10.1007/s40120-023-00546-w

[28] Guo J, Zhang R, Dong R, Yang F, Wang Y, Miao W. Interpretable machine learning model for predicting the prognosis of GBS patients. J Inflamm Res. 2024;17:5901–5913. doi:10.2147/JIR.S471626

[29] Guo J, Dong R, Zhang R, Yang F, Wang Y, Miao W. Interpretable machine learning model for predicting the prognosis of antibody positive autoimmune encephalitis patients. J Affect Disord. 2024;369:352–363. doi:10.1016/j.jad.2024.10.010

[30] Chicco D, Oneto L, Tavazzi E. Eleven quick tips for data cleaning and feature engineering. PLOS Comput Biol. 2022;18(12):e1010718. doi:10.1371/journal.pcbi.1010718

